# Experiences of testing negative or positive for HIV in Malawi, South Africa, and Zambia: a cross-sectional study

**DOI:** 10.1101/2025.03.24.25324300

**Authors:** N Mutanda, A Morgan, A Huber, N Scott, I Mokhele, T Tcherini, T Masina, R Nyirenda, A Kamanga, P Lumano-Mulenga, S Rosen, S Pascoe

## Abstract

**Background:** HIV testing services (HTS) aim to increase HIV status awareness and serve as the gateway to prevention, care, and treatment. Understanding clients’ experiences accessing facility-based HTS is important to ensure comprehensive service provision, improve linkage to care, and ultimately contribute to better health outcomes.

**Methods:** We surveyed a convenience sample of adults presenting for HIV testing at 42 health facilities in Malawi, South Africa, and Zambia between September 2022 and April 2023. A structured questionnaire captured data on HTS history, reasons for testing, services obtained, and offer and uptake of pre-exposure prophylaxis (PrEP) or treatment. Open-ended questions captured participants’ experiences qualitatively. We report participant characteristics, experiences, and services provided using proportions stratified by country and HIV test result and summarize emergent themes from qualitative responses.

**Results:** We enrolled 1,142 clients who presented for facility-based HIV testing (324 from Malawi, 389 from South Africa, 429 from Zambia). Of these, 32%, 24%, and 34% tested positive for HIV in Malawi, South Africa, and Zambia, respectively. Although most participants had tested for HIV prior to the current test, this proportion varied by country and current test result but was substantially lower for positive testers (64%, 66%, and 71% for Malawi, South Africa and Zambia) than for negative testers (82%, 88%, and 86%, respectively). Over a third of those who tested positive for HIV at this test had never previously tested for HIV. Among participants who tested positive, ill health was most frequently reported as the reason for testing; negative testers most often reported testing voluntarily (for their own information). Most participants reported being tested by lay counselors and receiving counseling before and after testing. Most participants who tested positive reported receiving ART initiation services, but fewer than a third in Malawi and South Africa and 39% in Zambia said they received adherence counselling. The proportion of those testing negative who were offered PrEP varied across the three countries, ranging from 27% of females in Malawi to 53% of females in Zambia. Participants in all three countries reported high satisfaction with the HTS services received and many qualitatively described high quality counseling, feelings of encouragement, and kind, warm treatment by providers.

**Conclusion:** While HIV testing clients in Malawi, South Africa, and Zambia were generally satisfied with their testing experiences, we identified opportunities for improvement in HTS services. ART adherence counseling was not universally provided, and prevention services were not offered to most negative testers. Of concern are the large proportion of positive testers who had never been tested before and self-reported testing because of ill health, which may indicate late presentation for testing. Emphasis on comprehensive linkage to services should be prioritized for all clients presenting for HIV testing.

**Statements and Declarations:** We have no financial or non-financial competing interests to disclose.

## Introduction

As the process of initiating treatment for HIV has been simplified and accelerated and options for HIV prevention have expanded in recent years, HIV testing has become a gateway both to immediate initiation of HIV treatment for those who test positive and access to effective prevention technologies for those who test negative [1]. The expansion of biomedical prevention options for HIV allows clients choices beyond condoms and voluntary male medical circumcision: in many settings, those testing negative now have access to both pre-exposure prophylaxis (PrEP) and post-exposure prophylaxis (PEP) [2]. With the advent of these new options, including the potential of long-acting medication formulations in the near future [3], the emphasis of HIV testing is now on promoting testing for both treatment and prevention and using a client-centered approach to engage all testers in appropriate follow up services, with those who test positive for HIV started on antiretroviral therapy (ART) and those who test negative offered prevention services [1,4].

Many countries have made progress toward the global target of 95% of individuals who are living with HIV being aware of their status, but in some countries a concerning minority remain undiagnosed. Linkage to treatment and prevention are also suboptimal. In Malawi and Zambia, the 2020-2021 Population Based HIV Impact Assessments (PHIA) found that 11.7% and 11.3% of adults were unaware of their HIV status, respectively [4,5]. South Africa estimates that 95% of people living with HIV (PWH) know their status [6], but only 77% are on ART, suggesting a gap in linkage to treatment. Data on linkage to prevention services for HIV-negative populations are scarce. While several estimates have been published of actual PrEP uptake (not offers of PrEP) among clients belonging to key populations (pregnant and post-partum women, sex workers, men who have sex with men) [7–10], we found only a single report of the proportion of the general population who test negative for HIV who are offered PrEP in routine care, from one district in South Africa [11].

To achieve the goals of linkage to treatment for those testing positive for HIV, linkage to prevention for HIV-negative testers, and optimizing prevention methods based on client risks and needs, it is important to adopt a client-centered approach and understand the characteristics, preferences, and recent testing experiences of people seeking HIV tests (HIV testers). We surveyed HIV testers at public healthcare facilities in Malawi, South Africa, and Zambia to describe who is testing for HIV now, to what degree testers are linked to services (regardless of testing outcome), and client acceptance of and views on treatment, prevention, and other supportive services.

## Methods

### Study sites and population

The AMBIT Project’s SENTINEL study [12] was conducted at 12 public sector clinics in Malawi, 18 in South Africa, and 12 in Zambia (Figure 1). The facilities were purposively chosen to ensure relatively large ART patient volumes, diversity of rural and urban settings, varied experience with differentiated service delivery (DSD) models for HIV treatment, and a range of HIV testing services (HTS). Additional information on the study sites is provided in the published protocol[12]. SENTINEL surveyed a sample of HIV testers between 16 September 2022 and 27 April 2023.

**Figure 1.**
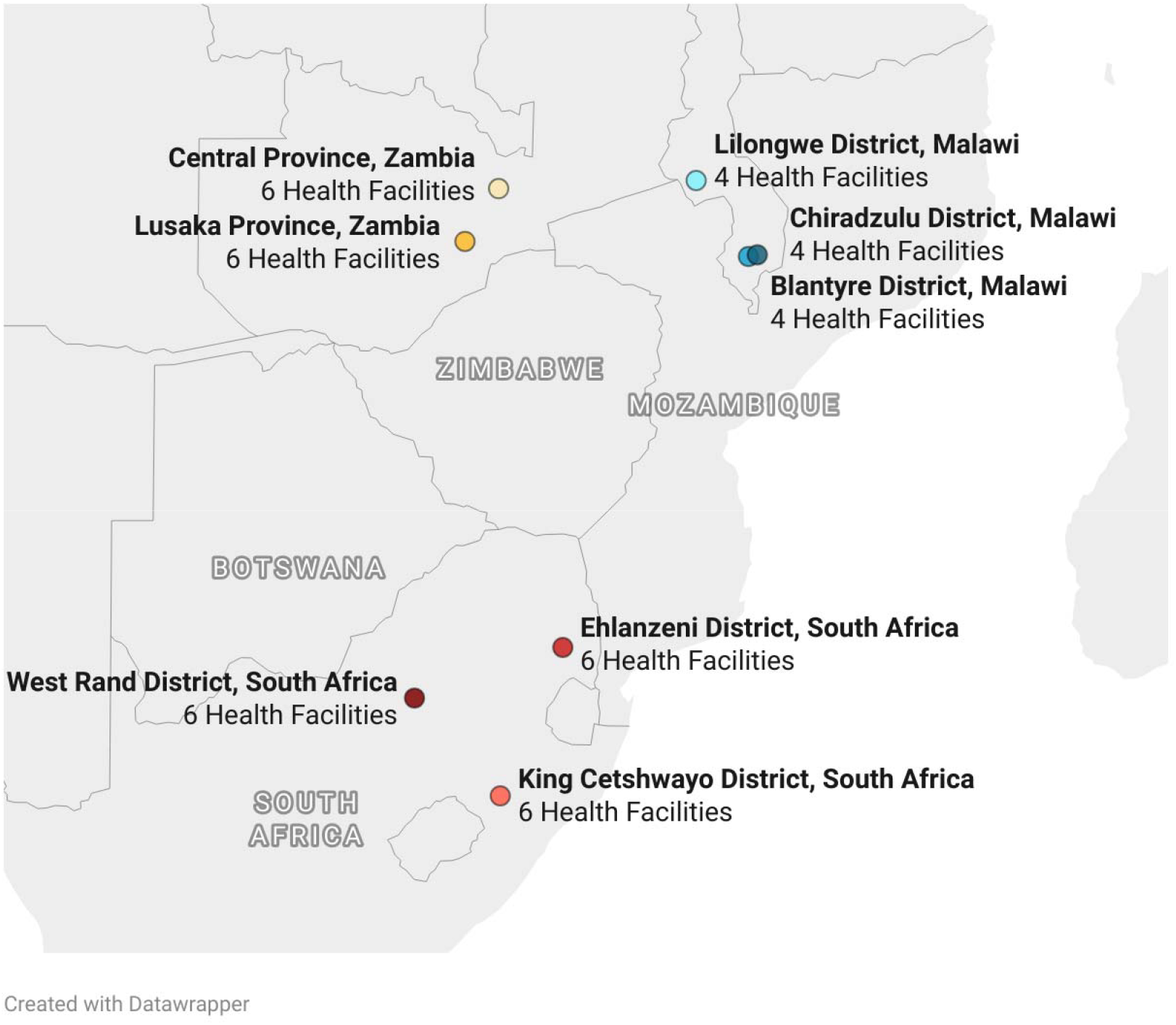
Study site locations across in the three study countries.

According to guidelines in all three countries, the HIV testing process should include voluntary consent, assurance of confidentiality, pre-test and post-test counselling, HIV testing, provision of accurate and complete test results, and effective linkage to prevention for those who test negative or linkage to treatment for those who test positive for HIV. HIV tests are to be performed by a trained and certified provider [13–15].

For the testing component of SENTINEL, we recruited a convenience sample of adults aged 16 years and above in Malawi and 18 years and above in South Africa and Zambia who presented for facility-based HIV testing. Primary healthcare clinics in the study countries typically offer HTS at several locations within the facility. In Zambia, HTS is offered in the HIV/ART, outpatient, and other departments, while in South Africa and Malawi, services are offered in HIV/ART, outpatient, antenatal care, maternity, paediatric care, HTS, chronic, acute, TB, and other consultation rooms. Clients can present voluntarily for testing through client-initiated counselling and testing, also called Voluntary Counselling and Testing (VCT). In all three countries, provider-initiated testing and counselling (PICT), under which a provider actively offers counselling and testing services to persons presenting at a health care facility for any reason as part of routine care, is also recommended, however adherence to this recommendation is unknown [13–15]. For this study we focused on clients who presented for HIV testing at health facility locations with HTS services through VCT and/or PICt. Clients testing at the antenatal care (ANC) clinic were excluded from the analysis as pregnant women generally comprise a different population with different procedures for linking to services.

At each site, we aimed to enroll up to 40 testers stratified by HIV status, for a maximum of 20 adults who tested positive for HIV (positive testers) and a maximum of 20 who tested negative (negative testers). Enrollment in the survey was not intended to create a representative sample of the overall population of HIV testers at the study sites, but rather to provide sufficient numbers of positive and negative testers to describe the experiences of each group. As a result, our results are neither intended to estimate nor imply proportions of clients testing positive or negative or the prevalence of characteristics among all testers. We thus stratify all results by country and HIV status and emphasize that findings are relevant only to the country/status group for whom they are reported.

### Data collection

Clients presenting for HIV testing were referred to a study research assistant by a member of the clinic staff either before or after their HIV test. The research assistant administered the informed consent process and documented written informed consent from eligible clients who agreed to participate. Those who consented either immediately responded to the survey (if they were approached after their HIV test) or completed testing, received their results and relevant services, and then responded to the survey. The survey included both quantitative questions and qualitative, open-ended questions. It captured HIV testing history, reason for testing on the day of study enrolment, location in the facility where the test was conducted, the department in the facility that the client had been referred from, and other services provided alongside testing. Self-reported results of the HIV test were recorded and verified using medical records. The questionnaire asked participants about the offer and acceptance of ART or of PrEP, according to their HIV test outcome. It also asked participants who tested negative for HIV whether other prevention strategies were discussed with the provider.

### Data analysis

We first calculated and present descriptive statistics, including frequencies and medians with interquartile ranges to describe study population characteristics, HIV testing experience, and offer/acceptance of ART or PrEP, stratified by HIV test outcome and country. We then conducted a content analysis of open-ended questions. Inductive coding was used to identify emergent themes. Codes were aggregated into larger categories as applicable [16]. Themes are presented stratified by country and test result. Illustrative quotes supporting the key themes are presented with client demographics, including self-reported satisfaction with testing services. Results were used to interpret and explain the quantitative findings.

### Ethics

Ethical approval to conduct this study was granted by University of Witwatersrand (Medical) Human Research Ethics Committee in South Africa (Protocol M210241), the National Health Science Research Committee (NHSRC) in Malawi (protocol 21/03/2672), ERES Converge Institutional Review Board in Zambia (Protocol 2021-Mar-012), and by the Boston University Medical Campus Institutional Review Board in the United States (Protocol H-41402). Data collectors were trained in research ethics, the overarching study, and the specific survey instrument. Written informed consent was obtained from each participant before the survey commenced.

## Results

### Study sample

Between 16 September 2022 and 27 April 2023, we enrolled 1,259 clients who presented for HIV testing at the SENTINEL sites across all three countries. We excluded from this analysis 117 clients who were enrolled in the survey while testing at ANC department, creating an analytic sample of 1,142 participants presenting for HIV testing at ART, chronic, TB, outpatient department, and HTS consultation rooms/departments. Of this sample, 344 participants tested positive for HIV (104 in Malawi, 92 in South Africa, 148 in Zambia) and 798 tested HIV-negative (220, 297, and 281, in Malawi, South Africa and Zambia respectively) (Table 1). Participants who tested positive for HIV were slightly older than negative testers. South African participants were more often female and more often unemployed than were those from the other countries. Most participants reported being unable to access a small amount of money ($5-6) for health care when needed and one third to one half of participants in each country reported experiencing food shortages. Characteristics are stratified by sex as well as status and country in Supplementary Table 1.

**Table 1.** Characteristics of study sample by country and HIV test result.

| Characteristic | Malawi N=324 |  | South Africa N=389 |  | Zambia N=429 |  |
| --- | --- | --- | --- | --- | --- | --- |
|  | Positive (N=104) | Negative (N=220) | Positive (N=92) | Negative (N=297) | Positive (N=148) | Negative (N=281) |
| Age (median Q1, Q3) | 30 (25, 41) | 28 (22, 35) | 31 (24, 40) | 26 (22, 34) | 33 (26, 41) | 26 (22, 32) |
| Female, n (%) | 56 (54) | 127 (58) | 65 (71) | 239 (80) | 95 (64) | 157 (56) |
| Marital status, n (%) |  |  |  |  |  |  |
| *Not married/no long-term partner* | 19 (18) | 52 (24) | 72 (78) | 256 (86) | 36 (24) | 123 (44) |
| *Married or long-term partner* | 53 (51) | 145 (66) | 16 (17) | 36 (12) | 69 (47) | 143 (51) |
| *Divorced/separated/widowed* | 32 (31) | 23 (10) | 4 (4) | 5 (2) | 43 (29) | 15 (5) |
| Highest level of education |  |  |  |  |  |  |
| *Primary or less* | 58 (56) | 97 (44) | 45 (49) | 78 (26) | 84 (57) | 111 (40) |
| *Secondary* | 40 (38) | 101 (46) | 38 (41) | 172 (58) | 57 (39) | 134 (48) |
| *Post-secondary* | 6 (6) | 22 (10) | 9 (10) | 47 (16) | 7 (5) | 36 (13) |
| Employment status |  |  |  |  |  |  |
| *Formal employment* | 15 (14) | 11 (5) | 19 (21) | 59 (20) | 11 (7) | 24 (9) |
| *Informal employment* | 63 (61) | 139 (63) | 20 (22) | 51 (17) | 92 (62) | 147 (52) |
| *Unemployed* | 23 (22) | 48 (22) | 44 (48) | 140 (47) | 41 (28) | 83 (30) |
| *Student/Trainee* | 3 (3) | 22 (10) | 9 (10) | 47 (16) | 4 (3) | 27 (10) |
| Food shortage within the household*, n (%) | 53 (50) | 88 (40) | 32 (35) | 73 (25) | 78 (53) | 137 (48) |
| Would have difficulty obtaining local equivalent of US\$5-6 for medical treatment, n (%) | 75 (72) | 151 (69) | 58 (63) | 146 (49) | 113 (76) | 203 (72) |

\*Food shortage within household defined as self-report that participant and/or people in their household go without food sometimes/often.

### HIV testing history and reasons for HIV test

Most participants in all three countries had tested for HIV prior to the test on the day of study enrollment, with proportions ranging from a low of 60% among male HIV-positive testers in Malawi to a high of 90% among male HIV-negative testers in South Africa (Table 2). Generally, women more often reported having taken two or more previous HIV tests than did men (Malawi 73% vs 61%, South Africa 91% vs 75% and Zambia 66% vs 60%). Some participants reported four or more previous tests, including 45% of negative testers in South Africa. A considerable proportion (36% in Malawi, 34% in South Africa, and 29% in Zambia) of those who tested positive, however, reported that they had never tested for HIV before their positive test on the day of study enrollment. Most participants indicated that they would prefer to receive HIV tests at the facility, rather than at community locations such as a local pharmacy or at home.

**Table 2:** Participants’ HIV testing history and reasons for testing on the day of enrollment by gender, HIV test result and country.

| Variable | Malawi |  |  |  |  |  | South Africa |  |  |  |  |  | Zambia |  |  |  |  |  |
| --- | --- | --- | --- | --- | --- | --- | --- | --- | --- | --- | --- | --- | --- | --- | --- | --- | --- | --- |
|  | Positive testers |  |  | Negative testers |  |  | Positive testers |  |  | Negative testers |  |  | Positive Testers |  |  | Negative Testers |  |  |
|  | Male<br>N=48 | Female<br>N=56 | Total<br>N=104 | Male<br>N=93 | Female<br>N=127 | Total<br>N=220 | Male<br>N=27 | Female<br>N=65 | Total<br>N=92 | Male<br>N=58 | Female<br>N=239 | Total<br>N=297 | Male<br>N=53 | Female<br>N=95 | Total<br>N=148 | Male<br>N=124 | Female<br>N=157 | Total<br>N=281 |
| Testing history |  |  |  |  |  |  |  |  |  |  |  |  |  |  |  |  |  |  |
| Ever tested for HIV,<br>n (%) | 29<br>(60) | 38<br>(68) | 67<br>(64) | 73<br>(78) | 107<br>(84) | 180<br>(82) | 15<br>(56) | 46<br>(71) | 61<br>(66) | 52<br>(90) | 209<br>(87) | 261<br>(88) | 40<br>(75) | 65<br>(68) | 105<br>(71) | 103<br>(83) | 140<br>(89) | 243<br>(86) |
| If yes, one time | 7<br>(24) | 10<br>(26) | 17<br>(25) | 23<br>(32) | 30<br>(28) | 53<br>(29) | 9<br>(60) | 7<br>(15) | 16<br>(26) | 8<br>(15) | 19<br>(9) | 27<br>(10) | 20<br>(50) | 34<br>(52) | 54<br>(51) | 37<br>(36) | 36<br>(26) | 73<br>(30) |
| If yes, two times | 15<br>(52) | 13<br>(34) | 28<br>(42) | 26<br>(36) | 36<br>(34) | 62<br>(34) | 3<br>(20) | 20<br>(43) | 23<br>(38) | 18<br>(35) | 49<br>(23) | 67<br>(26) | 9<br>(22) | 14<br>(22) | 23<br>(22) | 31<br>(30) | 56<br>(40) | 87<br>(36) |
| If yes, three times | 4<br>(14) | 9<br>(24) | 13<br>(19) | 9<br>(12) | 17<br>(16) | 26<br>(14) | - | 3<br>(7) | 3<br>(5) | 4<br>(8) | 45<br>(22) | 49<br>(19) | 6<br>(15) | 8<br>(12) | 14<br>(13) | 16<br>(16) | 19<br>(14) | 35<br>(14) |
| If yes, ≥ four times | 3<br>(10) | 6 (16) | 9 (13) | 15<br>(21) | 24 (22) | 39<br>(22) | 3<br>(20) | 16<br>(35) | 19<br>(31) | 22<br>(42) | 96<br>(46) | 118<br>(45) | 5<br>(12) | 9 (14) | 14 (13) | 19<br>(18) | 29 (21) | 48<br>(20) |
| Preferred HIV testing location, n (%) |  |  |  |  |  |  |  |  |  |  |  |  |  |  |  |  |  |  |
| Clinic | 45<br>(94) | 56<br>(100) | 101<br>(97) | 83<br>(89) | 117<br>(92) | 200<br>(91) | 26<br>(96) | 57 (88) | 83<br>(90) | 48<br>(83) | 197<br>(82) | 245<br>(82) | 51<br>(96) | 92 (97) | 143<br>(97) | 101<br>(81) | 142<br>(90) | 243<br>(86) |
| At home | 3 (6) | 0 (0) | 3 (3) | 7 (8) | 8 (6) | 15 (7) | 1 (4) | 8 (12) | 9<br>(10) | 6 (10) | 32 (13) | 38<br>(13) | - | - | - | 17<br>(14) | 12 (8) | 29<br>(10) |
| Other | - | - | - | 3 (3) | 2 (2) | 5 (2) | - | - | - | 4<br>(6) | 10<br>(4) | 14<br>(5) | 2<br>(4) | 3<br>(3) | 5<br>(4) | 6<br>(5) | 3<br>(2) | 9<br>(3) |
| Reasons for testing for HIV today, n (%) |  |  |  |  |  |  |  |  |  |  |  |  |  |  |  |  |  |  |
| Pregnancy | - | 11 (20) | 11 (11) | - | 23 (18) | 23<br>(10) | - | 8 (12) | 8 (9) | - | 109<br>(46) | 109<br>(37) | - | 3 (3) | 3 (2) | 4 (3) | 15 (10) | 19 (7) |
| Feeling ill | 21<br>(44) | 17<br>(30) | 38<br>(37) | 9<br>(10) | 23<br>(18) | 32<br>(15) | 15<br>(56) | 26<br>(40) | 41<br>(45) | 14<br>(24) | 25<br>(10) | 39<br>(13) | 25<br>(47) | 37 (39) | 62 (42) | 20<br>(16) | 32 (20) | 52<br>(19) |
| Partner/former<br>partner diagnosed<br>with HIV | 2 (4) | 2 (4) | 4 (4) | 2 (2) | 2 (2) | 4 (2) | 1 (4) | - | 1 (1) | 2 (3) | 1 (0) | 3 (1) | 1 (2) | 1 (1) | 2 (1) | 4 (3) | 3 (2) | 7 (2) |
| PrEP | - | - | - | 3 (3) | 1 (1) | 4 (2) | - | - | - | 5 (9) | 11 (5) | 16 (5) | - | - | - | 7 (6) | 6 (4) | 13 (5) |
| Checking status/<br>voluntary testing | 17<br>(35) | 18 (32) | 35 (34) | 67<br>(72) | 65 (51) | 132<br>(60) | 7 (26) | 25 (38) | 32<br>(35) | 34<br>(59) | 76 (32) | 110<br>(37) | 14<br>(26) | 37 (39) | 51 (34) | 74<br>(60) | 85 (54) | 159<br>(57) |
| Provider<br>recommendation | 7<br>(15) | 7 (12) | 14 (13) | 12<br>(13) | 12 (9) | 24<br>(11) | 3 (11) | 5 (8) | 8 (9) | 2 (3) | 17 (7) | 19 (6) | 13<br>(25) | 15 (16) | 28 (19) | 11 (9) | 14 (9) | 25 (9) |
| Other | 1 (2) | 1 (2) | 2 (2) | - | 1 (1) | 1 (0) | 1 (4) | 1 (2) | 2 (2) | 1 (2) | - | 1 (0) | - | 2 (2) | 2 (1) | 4 (3) | 2 (1) | 6 (2) |

Among men who tested positive, ill health was the most commonly cited reason for testing for HIV (44% Malawi, 56% SA, and 47% Zambia), followed by voluntary testing (Table 2). A similar pattern prevailed among women who tested positive for HIV in SA and Zambia, where more than a third of the women tested because of ill health (40% SA vs 39% Zambia) followed by voluntary testing (38% SA and 39% Zambia). These were also the most frequent reasons for testing among HIV-positive women in Malawi, but in reverse order: voluntary testing (32%) was slightly more common than feeling ill (30%).

Among men who tested negative, voluntary testing was most common, reported by roughly 3 out of 5 negative testers. While voluntary testing was also common among women, pregnancy was also cited as the reason for testing by 18% of female negative testers in Malawi, 10% in Zambia, and 46% in South Africa. Of those who tested negative, between 13 and 19% of participants indicated that feeling ill was their main reason for testing. Fewer than 10% of all clients who tested negative reported having a test in order to qualify for PrEP.

### Services offered to all HIV testers

Nearly all study participants in all three countries were tested at the health facility’s HTS and about 80% reported that their test was conducted by a lay counsellor (Table 3). In all three countries, about 10-20% of each subgroup said that they did not receive either pre- or post-test counseling. Fewer negative testers reported being offered post-test counseling than positive testers. Offers of condoms and family planning were also less common, ranging from 14% to 38% for offers of condoms and 5% to 18% for family planning. Nearly half of the respondents noted that they were not provided with a confidential space for counseling.

### Services offered to positive testers

The majority of participants who tested positive for HIV were offered the opportunity to start ART on the day of testing: over 90% of participants who tested positive in Malawi and Zambia (including 100% of women in Malawi) were offered ART. 81% of men and 92% of women who tested positive in South Africa were also offered ART (Table 4). Of those who were offered, over 80% accepted the offer to start ART. Fewer than a quarter of participants in Malawi (14/104, 13%) and South Africa (20/92, 21%) reported being offered ART adherence counselling, and only just over a third of participants in Zambia received adherence counseling. Participant reports for having a CD4 test performed, varied widely across participant groups: from 21% and 29% of female and male participants in Malawi, to 67% and 40% of male and female participants in South Africa.

**Table 3:** Self-reported services offered to all HIV testers by HIV test results in Malawi, South Africa and Zambia.

|  | Malawi |  | South Africa |  | Zambia |  |
| --- | --- | --- | --- | --- | --- | --- |
|  | Positive<br>(N=104) | Negative<br>(N=220) | Positive<br>(N=92) | Negative<br>(N=297) | Positive<br>(N=148) | Negative<br>(N=281) |
| Location of today's HIV test |  |  |  |  |  |  |
| In-facility: HTS* | 103 (99) | 216 (98) | 90 (98) | 287 (97) | 120 (81) | 231 (82) |
| Other departments** | 1 (1) | 4 (2) | 2 (2) | 10 (3) | 28 (19) | 50 (18) |
| Who conducted your HIV test? |  |  |  |  |  |  |
| Nurse | 20 (19) | 50 (23) | 15 (16) | 42 (14) | 22 (15) | 36 (13) |
| Lay counsellor | 83 (80) | 169 (77) | 77 (84) | 253 (85) | 123 (83) | 232 (83) |
| Other (specify) | 1 (1) | 1 (0) | - | 2 (1) | 3 (2) | 13 (5) |
| Services received at today's visit |  |  |  |  |  |  |
| Pre-test counselling | 93 (89) | 179 (81) | 89 (97) | 264 (89) | 137 (93) | 238 (85) |
| Post-test counselling | 96 (92) | 182 (83) | 88 (96) | 231 (78) | 144 (97) | 225 (80) |
| Confidential space | 64 (62) | 116 (53) | 53 (58) | 164 (55) | 92 (62) | 140 (50) |
| Condoms | 28 (27) | 53 (24) | 35 (38) | 103 (35) | 20 (14) | 48 (17) |
| Family planning*** | 8 (8) | 39 (18) | 14 (15) | 50 (17) | 7 (5) | 17 (6) |
| TB test | 11 (11) | 4 (2) | 32 (35) | 7 (2) | 14 (9) | 1 (0) |

\* HIV testing service (HTS)
\*\* Other departments included outpatient department (OPD), ART clinic
\*\*\*Family planning services were offered to men less often than women: Among men who tested positive, services were offered to 1% in Malawi, 0% South Africa, and 1% in Zambia. Among men to tested negative, services were offered to 11% of men in Malawi, 2% in South Africa, and 2% in Zambia.

**Table 4:** ART initiation for participants who tested HIV-positive, by country and sex.

| Variable | Malawi |  | South Africa |  | Zambia |  |
| --- | --- | --- | --- | --- | --- | --- |
|  | Male<br>(N=48) | Female<br>(N=56) | Male<br>(N=27) | Female<br>(N=65) | Male<br>(N=53) | Female<br>(n=95) |
| Offered the chance to start or restart ART, n (%) | 46 (96) | 56 (100) | 22 (81) | 60 (92) | 48 (91) | 87 (92) |
| CD4 test performed | 14 (29) | 12 (21) | 18 (67) | 26 (40) | 10 (19) | 32 (34) |
| If offered, accepted offer to start or restart ART, n (%) | 46 (100) | 55 (98) | 21 (95) | 57 (95) | 39 (81) | 74 (85) |
| Offered adherence counselling | 8 (17) | 6 (11) | 5 (19) | 15 (23) | 17 (32) | 41 (43) |
| Main reason for starting or restarting ART, n (%) |  |  |  |  |  |  |
| Pregnancy | 0 (0) | 10 (18) | 0 (0) | 7 (12) | 0 (0) | 3 (3) |
| Feeling ill and wanted to feel better | 21 (46) | 12 (21) | 10 (45) | 20 (33) | 23 (48) | 33 (38) |
| Health care provider recommendation | 19 (41) | 29 (52) | 8 (36) | 19 (32) | 9 (19) | 19 (22) |
| Was worried about not being on treatment | 4 (9) | 4 (7) | 3 (14) | 6 (10) | 9 (19) | 17 (20) |
| Other | 2 (4) | 1 (2) | 1 (5) | 8 (13) | 7 (15) | 15 (17) |

In South Africa and Zambia for both males and females, ill health and wanting to feel better was the most commonly reported reason for same-day ART initiation, followed by health care provider recommendation. In Malawi, 52% of females cited health care provider recommendation as the main reason for starting ART that day followed by ill health (21%); among males the most cited reason was ill health and wanting to feel better (46%) followed by health care provider recommendation (41%) (Table 4).

Qualitative findings reinforced the quantitative results, suggesting that clients accepted ART because they wanted to feel better and desired to prioritize their health, as illustrated by the quotes below.

> “I have been feeling ill so I know this medication will make me start feeling better.” *– Male, age 25-29 years, Malawi*
>
> “My health is deteriorating and lost too much weight and decided for me to get better I need to start immediately” *-Male, age 45-49 years, South Africa*
>
> “I accepted so that I can continue living a normal life and see my children grow” -*Female, age 45-49 years, Zambia*

Of the 319 clients who were offered the chance to start ART on the day of testing, 27 refused medication. Most of these clients (n=22) were from Zambia and reported that they were not ready to start, needed time to process the information or inform family members, and described concerns over stigma:

> “I am not ready to start. I need to digest the results and tell my mum about them, then I’ll come back.” *-Female, age < 20 years, Zambia*
>
> “Because I would love to initiate from another facility away from this one because of stigma.” -*Female, age 45-49 years, Zambia*
>
> “I need to come with my wife for a test tomorrow so we can start together” -*Male, age 35-39 years, Zambia*

### Services offered to negative testers

As shown in Table 5, between 10% and 18% of clients reported that they did not learn as much as they would have liked about HIV and HIV prevention. Among negative testers, 28% in Malawi, 47% in South Africa, and 50% in Zambia were offered PrEP. PrEP acceptance among those offered PrEP varied from a minimum of 42% of men in Zambia to a maximum of 82% of women in Malawi.

**Table 5:** Prevention information and PrEP offer among those testing negative, by country and sex.

| Variable | Malawi |  | South Africa |  | Zambia |  |
| --- | --- | --- | --- | --- | --- | --- |
|  | Male<br>(N=93) | Female<br>(N=127) | Male<br>(N=58) | Female<br>(N=239) | Male<br>(N=124) | Female<br>(N=157) |
| Learned as much about HIV and HIV prevention as desired, n (%) | 84 (90) | 113 (89) | 50 (86) | 211 (88) | 102 (82) | 142 (90) |
| Offered PrEP, n (%) | 27 (29) | 34 (27) | 27 (47) | 114 (48) | 57 (46) | 83 (53) |
| If offered, accepted PrEP, n (%) | 19 (70) | 28 (82) | 18 (66) | 61 (54) | 24 (42) | 38 (46) |

For those who did not accept the offer of PrEP, reasons for rejecting the offer varied by country (Table 6). Emergent themes centred around risk perception, readiness, insufficient knowledge, and feeling as if they did not want or need it.

**Table 6.** Reasons for rejecting PrEP offer among those who tested negative, by country.

| Malawi (n=12) | South Africa* (n=62) | Zambia* (n=78) |
| --- | --- | --- |
| *Themes* |  |  |
| No concerns regarding HIV exposure/no perceived feeling of HIV risk<br>Feelings of unease, uncertainty, and risk associated with PREP and its side effects | Clients were undecided or not yet ready to start PREP; needed time to think about it<br>High burden/discomfort of taking daily medication<br>Pregnancy related concerns; willingness to initiate PREP after delivery<br>Not sexually active/no partner makes client feel at low risk of exposure | Lacking knowledge or need more information about PREP<br>No concerns regarding HIV exposure/no perceived feeling of HIV risk<br>Negative HIV status, possibly pointing to misconceptions about PREP use |
| *Illustrative quotes* |  |  |
| "Not a requirement and [I am] not at risk" -Male, age <20 years | "I will come back soon to initiate" -Female, age 25-29 years | "I don't understand what PrEP is" -Male, age <20 years |
| "I don't know how it works" -Female, age 20-24 years | "I don't want to take tablets every single day of my life" -Female, age <20 years | "Because we were guided that its mostly given to those who are positive." -Male, age 20-24 years |
|  | "Might opt to start post-delivery of the baby" -Female, age 35-39 years | "I'm not in a relationship with an infected person" -Female, age 20-24 years |

\*9/62 clients in South Africa and 36/78 clients in Zambia noted that they did not like, did not want, or did not need PrEP but did not expand on the specific deterrents.

### Client satisfaction and recommendations to improve testing

Over 90% of the participants of both sexes and of those testing positive and negative in Malawi and Zambia reported that they were satisfied with the testing service they received. Proportions reporting satisfaction with testing services were almost as high in South Africa, exceeding 90% for HIV-negative testers of both sexes and reaching 89% for HIV-positive testers.

Nearly all (98%) clients reported that the care they received was welcoming and supportive, and clients in all three countries qualitatively attributed positive provider disposition and feeling welcome in the facility to their overall satisfaction. Clients described providers as friendly, uplifting, and engaging and that the clinic was welcoming. Many clients reported that their providers helped subdue fears about testing and treatment initiation, treating clients with kindness and dignity throughout the testing experience.

> “The doctor explained everything and convinced me that it is not the end of the world and I need to take heart. So, I am happy despite the status*” – Female, age 35-39 years, very satisfied with care on day of HIV testing, tested positive for HIV, Malawi*
>
> “The counselor was calm explaining and giving information on all I needed to know, she sounded experienced” *– Female, age 25-29 years, satisfied with care on day of HIV testing, tested negative for HIV, South Afric*a
>
> “I really didn’t expect the warm welcome I got. The providers were very kind and helpful*” – Male, age 45-49 years, very satisfied with care on day of HIV testing, tested positive for HIV, Zambia*

Clients also reported positive experiences and satisfaction with the counseling they received.

> “I was very happy with the care I received today and they prepared me very well before hearing my results…hence I was all ready for any outcome” *– Male, age 30-34 years, satisfied with care on day of HIV testing, tested negative for HIV, Malawi*
>
> “It was easy for me to communicate with the health counsellor. She told me everything I wanted to know about HIV.” *– Female, age 25-29 years, satisfied with care on day of HIV testing, tested negative for HIV, South Africa*
>
> “I feel encouraged and free to open up to them and ask anything.” *– female, age 45-49 years, satisfied with care on day of HIV testing, tested positive for HIV, Zambia*

Many clients reported that they were satisfied that they now knew their status, acknowledging that it was important to navigate their health. Others expressed emotional responses of relief, confusion, or fear regarding the test outcome. The few who reported a negative experience explained that they experienced long waiting times, long queues, slow clinic operations, and internal anxiety about testing.

To improve services, participants primarily recommended more staff and shorter wait times, followed by more information and counselling (Table 7).

**Table 7.** Self-reported client satisfaction and recommended service improvements, by country, sex, and HIV status.

| Variable | Malawi |  |  |  | South Africa |  |  |  | Zambia |  |  |  |
| --- | --- | --- | --- | --- | --- | --- | --- | --- | --- | --- | --- | --- |
|  | Male |  | Female |  | Male |  | Female |  | Male |  | Female |  |
|  | HIV+ | HIV- | HIV+ | HIV- | HIV+ | HIV- | HIV+ | HIV- | HIV+ | HIV- | HIV+ | HIV- |
| N | 48 | 93 | 56 | 27 | 27 | 58 | 65 | 23 | 53 | 124 | 95 | 157 |
| Satisfied with care received during HIV test*, n (%) | 44 (92) | 84 (91) | 52 (93) | 121 (95) | 24 (89) | 53 (91) | 58 (89) | 222 (93) | 48 (91) | 112 (90) | 87 (92) | 151 (96) |
| Need for service improvement, n (%) |  |  |  |  |  |  |  |  |  |  |  |  |
| More staff | 14 (29) | 35 (38) | 18 (32) | 57 (45) | 10 (37) | 14 (24) | 30 (46) | 102 (43) | 14 (26) | 29 (23) | 27 (28) | 34 (22) |
| More information provided by staff | 6 (12) | 12 (13) | 5 (9) | 24 (19) | 1 (4) | 3 (5) | 3 (5) | 26 (11) | 6 (11) | 6 (5) | 9 (9) | 12 (8) |
| Shorter waiting time | 29 (60) | 33 (35) | 39 (70) | 55 (43) | 14 (52) | 21 (36) | 30 (46) | 89 (37) | 14 (26) | 27 (22) | 18 (19) | 37 (24) |
| More counselling when there are problems | 4 (8) | 11 (12) | 6 (11) | 12 (9) | 1 (4) | 10 (17) | 9 (14) | 26 (11) | 8 (15) | 7 (6) | 9 (9) | 13 (8) |
| More counselling overall | 2 (4) | 7 (8) | 2 (4) | 7 (6) | 5 (19) | 7 (12) | 11 (17) | 42 (18) | 8 (15) | 13 (10) | 14 (15) | 17 (11) |

\*Asked using a Likert scale, where clients were very satisfied, satisfied, neither satisfied nor dissatisfied, dissatisfied, or very dissatisfied with care. Satisfaction in this case refers to all clients who responded that they were satisfied or very satisfied.

## Discussion

This study aimed to understand the experiences of clients who present for HIV testing at health care facilities in Malawi, South Africa, and Zambia in the era of client-centered HIV testing. We found that reasons for testing and experiences vary by HIV test result. The most common reason for HIV testing among those who tested positive was feeling unwell, whereas among those who tested negative, the most frequently cited reason was self-initiated voluntary testing to check their status. Clients preferred facility-based HIV testing over other community testing locations such as pharmacies and homes. Across all three countries, nearly all positive testers were offered ART and nearly all accepted ART. Only about a third to a half of negative testers were offered PrEP. Clients in all three countries were satisfied with their testing experience, though many wanted to see more staffing and shorter waiting times, and some expressed a desire for more counselling.

In this era of testing for treatment and prevention, our study elicited both encouraging and concerning findings. We were encouraged to find near-universal satisfaction with HTS and providers, in view of recent literature suggesting that clinic obstacles and provider attitudes are common reasons for disengagement from care [17,18]. Most of these sources pertain to HIV treatment rather than testing, however, it may be that problems of provider disposition do not pertain to testing, or that the role of counselors, who remain central to HTS, mitigates negative experiences with other providers. Although a majority of participants suggested shorter waiting times as a desired service improvement, long waits did not seem to diminish their satisfaction with the service received. While we didn’t ask clients about their expectations for care, some illustrative quotes suggest that clients were surprised by the good quality of care that they received. It is possible that this misaligned expectation impacts overall satisfaction – while some things were unsatisfactory (wait times), their overall experience at the clinic was better than expected.

It was also encouraging to observe that nearly all HIV-positive testers were both offered and accepted immediate treatment initiation, suggesting that “linkage to care” between testing and treatment is no longer a major point of loss in the care and treatment cascade. We note that our sample does not represent all positive testers, because those who are too ill for primary healthcare clinics will go directly to hospitals, and opportunistic infections may delay ART initiation for some of these patients. We also observe that a significant proportion of ART initiators--9.4% in South Africa and 21% in Zambia in two recent studies [19,20]--never return to the clinic after the visit at which they initiated. That said, our data show that the effort to simplify and accelerate ART initiation over the past decade appears to have been successful.

On the concerning side was our finding that roughly one third of positive testers in all three countries said that they have never had an HIV test before their positive test on the day of study enrollment. Interestingly, negative testers of both sexes and in all three countries were more likely to have prior testing experience than were those who tested positive. Since HIV testing is presumably no longer conducted after a positive test, it may be anticipated that negative testers receive more tests over a lifetime. The fact that negative testers in our sample were slightly younger than positive testers, however, suggests that the positive testers were less likely to seek diagnosis overall until ill health drove them to take action, another point of concern.

Related to our first concern is that so many respondents in all three countries reported testing because of ill health. Despite efforts to destigmatize HIV and promote status neutral testing, it appears that many individuals still regard HIV testing as an action only to be taken when ill, rather than a regular preventative measure, and that many are not aware of their own HIV infection risk. Existing evidence from the region indicates that late presentation for HIV care and advanced HIV disease (AHD) at initiation remain a problem [21,22]. AHD is harmful to patients, whose prognosis is worse than for early starters; challenging for providers, as cases are typically more complicated and require careful monitoring [23,24]; and threatening to public health efforts to suppress community viral load [25].

A third serious concern pertains to the low proportion of HIV testers who were offered prevention options. Only one out of six participants self-reported being offered condoms in Zambia, a rate that rose to only just over 30% in South Africa. There was little difference between positive and negative testers in their report of condom offers. Similarly, fewer than half of negative testers said they were offered PrEP, with the exception of female testers in Zambia, of whom 53% indicated being offered PrEP. Interestingly, acceptance of the offer of PrEP was highest in Malawi, where the likelihood of offer itself was lowest. This phenomenon may reflect selection on the part of providers, who may be more likely to offer PrEP to those they believe will accept it. Relatively low rates of acceptance in Zambia - fewer than half of those offered PrEP accepted it - may merit further investigation in that country.

While PrEP availability is increasing throughout sub-Saharan Africa, relative to other prevention methods and to ART for HIV treatment, it is still quite novel. A growing body of behavioral research, primarily qualitative, suggests that obstacles to PrEP uptake by HIV-negative persons are similar to those for ART uptake among HIV-positive individuals. In Zambia, for example, women and other key populations have recently reported needing partner approval, having negative experiences with the health provider, anticipated stigma or being mistaken for having HIV, distance to clinic, and clinic waiting times as barriers to PrEP uptake [26,27]. In South Africa, reported barriers have included limited knowledge or awareness of PrEP [28], stigma and partner approval among pregnant and pre-conception women [29], and fear, stigma, partner support, clinic wait times, and health provider attitudes as barriers among young people [30,31]. Our results, combined with findings from other studies, suggest a need for improved information, education, and communication support strategies to address knowledge and concerns and build trust in PrEP to ensure all those eligible can benefit.

Finally, we noted that study respondents felt strongly about counseling. Quality and desire for counseling emerged as themes throughout the survey. When all testers were asked about the worst part of the testing experience, many described being nervous, emotional, and afraid about test results; however, they also described that compassionate and comprehensive counseling helped to ease their fears and improve the experience. Most clients in all countries said they received both pre- and post-test counseling, as called for by guidelines. There was a gap in post-test counseling, though, with some 20% of negative testers reporting that they did not receive post-test counseling. Despite this, most clients who tested negative said that they learned as much about HIV prevention as they wanted and describe positive experiences with counseling.

Of concern is that only between 13% (Malawi) and 39% (Zambia) of HIV-positive testers reported receiving adherence counselling as part of routine care after their positive test. Adherence counseling following a positive test and/or ART initiation is called for in all countries’ HTS guidelines [32–34]. While little is known about its effectiveness in improving treatment outcomes, it is likely to be beneficial to at least some patients, and failure to provide it warrants health system attention. Previous studies have described inadequate counseling at HIV testing clinics leaving clients feeling unprepared and with unresolved emotions about their diagnosis, underscoring the importance of comprehensive client-centered care [35].

Our study had a number of limitations. Although we intended to enroll a sample of testers representative of the negative and positive populations who are seeking HIV tests at the study sites, the large number of testing locations within clinics (e.g. HIV clinic, outpatient clinic, TB clinic, etc.), inconsistent referral to our research assistants by clinic staff, and generally small number of positive testers led to enrollment of a convenience sample, rather than a truly representative sample. As such, our results cannot be interpreted as indicating the proportion of negative and positive testers at the site. We believe that our results do reflect the experiences of clients testing positive or negative at the study facilities’ HTS locations at the time of the study. A second limitation pertains to the self-reported nature of our data. Questions about specific services may have caused confusion among some respondents, leading to unreliable answers. It is possible, for example - though solely speculation on our part - that some participants confused adherence counselling with post-test counselling, and thus reported not having adherence counselling. We also relied on clients’ self-reported accounts of offer and acceptance of PrEP and were unable to confirm these accounts, which could produce either under- or over-estimates of PrEP utilization. Finally, our overall sample was small in terms of both numbers of facilities and numbers of testers at each facility. While we have no reason to suspect that our results are not reflective of clients’ characteristics nationally, generalizing beyond the study sites should be done with caution.

## Conclusion

In 2023, more than 90% of people living with HIV in eastern and southern Africa were aware of their HIV status, according to UNAIDS [36]. Access to HIV testing is thus likely nearly universal in these regions. In this study of the experience of positive and negative testers in Malawi, South Africa, and Zambia, we found that nearly all of them were satisfied with their experience of testing and appreciative of the services they received. At the same time, our results showed that a large proportion of positive testers had never been tested before and that ill health remained the most common reason for seeking an HIV test, both indications of late testing. We also found that while same-day treatment initiation was offered to nearly all who tested positive, ART adherence counseling was not consistently provided. Among those who tested negative, moreover, only a minority were offered HIV prevention strategies like PrEP. We conclude that while current facility-based HIV testing provides a generally satisfactory experience to clients, there remains substantial room for improvement in the quality and completeness of the services offered.

## Data Availability

All data produced in the present study are available upon reasonable request to the authors

---

**Supplementary Table 1:** Participants characteristics by gender, HIV test result and Country.

|  | Malawi |  |  |  | South Africa |  |  |  | Zambia |  |  |  |
| --- | --- | --- | --- | --- | --- | --- | --- | --- | --- | --- | --- | --- |
|  | Male |  | Female |  | Male |  | Female |  | Male |  | Female |  |
|  | Positive<br>(N = 48) | Negative<br>(N = 93) | Positive<br>(N = 56) | Negative<br>(N = 127) | Positive<br>(N = 27) | Negative<br>(N = 58) | Positive<br>(N = 65) | Negative<br>(N = 239) | Positive<br>(N = 53) | Negative<br>(N = 124) | Positive<br>(N = 95) | Negative<br>(N = 157) |
| **Age, median (IQR)** | 31<br>(25, 43) | 28<br>(24, 33) | 30<br>(24, 40) | 27<br>(21, 35) | 39<br>(31, 47) | 32<br>(25, 40) | 30<br>(24, 37) | 25<br>(22, 32) | 36<br>(31, 42) | 25<br>(22, 30) | 29<br>(24, 40) | 26<br>(22, 33) |
| **Marital status, n (%)** |  |  |  |  |  |  |  |  |  |  |  |  |
| *Never married* | 13 (27) | 26 (28) | 6 (11) | 26 (20) | 15 (56) | 44 (76) | 57 (88) | 212 (89) | 9 (17) | 67 (54) | 27 (28) | 56 (36) |
| *Married* | 20 (42) | 60 (65) | 33 (59) | 85 (67) | 9 (33) | 13 (22) | 7 (11) | 23 (10) | 28 (53) | 56 (45) | 41 (43) | 87 (55) |
| *Divorced/separated/widowed* | 15 (31) | 7 (8) | 17 (30) | 16 (13) | 3 (11) | 1 (2) | 1 (2) | 4 (2) | 16 (30) | 1 (1) | 27 (28) | 14 (9) |
| **Highest level of education, n (%)** |  |  |  |  |  |  |  |  |  |  |  |  |
| *No schooling/primary* | 27 (56) | 39 (42) | 31 (55) | 58 (46) | 17 (63) | 28 (48) | 28 (43) | 50 (21) | 27 (51) | 59 (48) | 57 (60) | 52 (33) |
| *Secondary* | 20 (42) | 45 (48) | 20 (36) | 56 (44) | 8 (30) | 23 (40) | 30 (46) | 149 (62) | 25 (47) | 50 (40) | 32 (34) | 84 (54) |
| *Post-secondary* | 1 (2) | 9 (10) | 5 (9) | 13 (10) | 2 (7) | 7 (12) | 7 (11) | 40 (17) | 1 (2) | 15 (12) | 6 (6) | 21 (13) |
| **Employment status, n (%)** |  |  |  |  |  |  |  |  |  |  |  |  |
| *Formal employment* | 8 (17) | 7 (8) | 7 (12) | 4 (3) | 8 (30) | 21 (36) | 11 (17) | 38 (16) | 7 (13) | 11 (9) | 4 (4) | 13 (8) |
| *Informal employment* | 32 (67) | 62 (67) | 31 (55) | 77 (61) | 12 (44) | 18 (31) | 8 (12) | 33 (14) | 45 (85) | 75 (60) | 47 (49) | 72 (46) |
| *Unemployed* | 5 (10) | 12 (13) | 18 (32) | 36 (28) | 6 (22) | 16 (28) | 38 (58) | 124 (52) | 1 (2) | 30 (24) | 40 (42) | 53 (34) |
| *Student/Trainee* | 3 (6) | 12 (13) | 0 (0) | 10 (8) | 1 (4) | 3 (5) | 8 (12) | 44 (18) | 0 (0) | 8 (6) | 4 (4) | 19 (12) |
| **Food scarcity, n (%)** |  |  |  |  |  |  |  |  |  |  |  |  |
| *Never* | 18 (38) | 28 (30) | 19 (34) | 53 (42) | 14 (52) | 41 (71) | 38 (58) | 155 (65) | 12 (23) | 41 (33) | 41 (43) | 69 (44) |
| *Seldom* | 8 (17) | 26 (28) | 6 (11) | 25 (20) | 3 (11) | 6 (10) | 5 (8) | 22 (9) | 7 (13) | 23 (19) | 10 (11) | 11 (7) |
| *Sometimes* | 19 (40) | 33 (35) | 21 (38) | 35 (28) | 7 (26) | 10 (17) | 13 (20) | 57 (24) | 32 (60) | 56 (45) | 39 (41) | 66 (42) |
| *Often* | 3 (6) | 6 (6) | 10 (18) | 14 (11) | 3 (11) | 1 (2) | 9 (14) | 5 (2) | 2 (4) | 4 (3) | 5 (5) | 11 (7) |
| **Would have difficulty obtaining R 100/1000 Kwacha for medical treatment, n (%)** | 37 (77) | 63 (67) | 38 (68) | 88 (69) | 18 (67) | 28 (48) | 40 (62) | 118 (49) | 38 (72) | 86 (69) | 75 (79) | 117 (75) |

## Notes

### Competing Interest Statement

The authors have declared no competing interest.

### Clinical Trial

NCT05886530

### Funding Statement

Funding for the study was provided by the Bill & Melinda Gates Foundation through OPP1192640 to Boston University and INV-037138 to the Wits Health Consortium.

### Author Declarations

Ethical approval to conduct this study was granted by University of Witwatersrand (Medical) Human Research Ethics Committee in South Africa (Protocol M210241), the National Health Science Research Committee (NHSRC) in Malawi (protocol 21/03/2672), ERES Converge Institutional Review Board in Zambia (Protocol 2021-Mar-012), and by the Boston University Medical Campus Institutional Review Board in the United States (Protocol H-41402).

